# Improvement in Expanded Disability Status Scale (EDSS) and anti-inflammatory parameters in patients with multiple sclerosis following oral consumption of N-163 strain of *Aureobasidium pullulans* produced beta glucan in a pilot clinical study

**DOI:** 10.1101/2023.05.14.23289953

**Authors:** Vidyasagar Devaprasad Dedeepiya, Chockanathan Vetrievel, Nobunao Ikewaki, Koji Ichiyama, Naoki Yamamoto, Hiroto Kawashima, Sudhakar S Bharatidasan, Subramaniam Srinivasan, Rajappa Senthilkumar, Senthilkumar Preethy, Samuel JK Abraham

## Abstract

**Introduction:** Multiple Sclerosis (MS) is a debilitating neurodegenerative disease in which demyelination due to auto-inflammation is considered to be the underlying pathogenesis, though the exact etiology is not known. Most of the management strategies involve medications that are anti-inflammatory or immune-suppressive, which do have associated side effects. In this study we have evaluated in MS patients, the clinical effects of a novel beta-glucan which has a track record of anti-inflammatory, immune-modulating potentials in earlier clinical and pre-clinical studies.

**Method:** The study involved 12 MS patients who consumed two sachets of N-163 strain of *Aureobasidium pullulans* produced B-Glucan, daily for 60 days along with routine medication.

**Results:** The Expanded Disability Status Scale (EDSS) improved by 0.5 in two patients and by 1 in one patient post-intervention, worsened in 1 patient, remaining stable in the rest. Decrease in IL-6, improvement in CD4+ve, CD19+ve, CD3+ve, and CD8+ ve cell count, increase in Lymphocyte to C-reactive protein ratio (LCR), Leukocyte to CRP ratio (LeCR) and a decrease in Neutrophil to Lymphocyte ratio (NLR) were observed.

**Conclusion:** This study having proven the safety of N-163 strain of *A*.*pullulans* produced B-Glucan food supplement and the efficacy by improvement in the EDSS score, besides beneficial modulation of inflammation and immune parameters of relevance in MS patients in a short duration of 60 days, has significant potential as a disease modifying adjuvant in MS. Immunological parameters like NLR, LCR, LeCR correlating with clinical improvement, in line with earlier reports using the same beta-glucans, gain further significance for their potentials as biomarkers in MS.

## Introduction

Multiple Sclerosis (MS) is an autoimmune illness of the central nervous system with a prevalence of more than 2 million people globally. MS is a diverse illness, without a specific etiology, but several factors are considered to play a role including environmental and genetic factors such as the relationship between HLA-DRB1*15:01 and vitamin D status, obesity, smoking, and infection with the Epstein-Barr virus (EBV). MS results in significant physical or cognitive impairment as well as neurological issues, especially in young adults [1]. Pathogenesis starts with the destruction of myelin sheath that leads to the formation of CNS plaques made up of inflammatory cells and their by-products, demyelinated and transected axons, and astrogliosis in both white and grey matter. The underlying mechanism involves focal T-lymphocytic and macrophage infiltrations, as well as death of oligodendrocytes. These lesions jeopardize the nerve impulse transmission and cause neuronal dysfunction, including autonomic and sensory deficits, visual disturbances, ataxia, weariness, and emotional issues [1].

Sub-types of MS are relapsing remitting MS (RRMS), primary progressive MS (PPMS), secondary progressive MS (SPMS), and progressive relapsing MS (PRMS). MS pathogenesis is influenced by changes in the peripheral immune system, blood brain barrier permeability, and intrinsic CNS immune cells (such as microglia) [2]. These three aspects of MS pathogenesis are the focus of current therapy approaches. Neurodegeneration, acute and chronic inflammation occur throughout the course of the disease, with acute inflammation being more prominent during the relapsing phase of the illness. There is no definite focus for a therapeutic approach to MS due to the disease’s diversity and hence currently, no remedy.

Current disease-modifying therapies (DMT)s’ primary objective is to keep the condition under control by lowering inflammation, myelin damage, and relapses [2]. Early “Traditional” Injectable DMTs include Interferon Beta and Glatiramer Acetate. Then came medications such as Sphingosine-1-Phosphate (S1P) Receptor Modulators, Teriflunomide, Fumarates and Cladribine. Infusion and injectable DMTs are monoclonal antibodies such as Natalizumab, Alemtuzumab, Rituximab, Ocrelizumab and Ofatumumab. Oral medications under investigation include Bruton’s Tyrosine Kinase Inhibitors (BTKi). An emerging new therapy is autologous hematopoietic stem cell transplantation (AHSCT) which aims to reset the immune system after existing autoreactive immune cells are eliminated with either fully or partially myeloablative conditioning. All the above mentioned therapies are associated with side effects such as gastrointestinal (GI) problems, flushing, severe liver injury, macular edema, bradycardia and increased risk for malignancy [2,3] The heterogeneous nature of the disease, which is impacted by environmental and genetic variables as well as the immune system’s naturally adaptive and changing nature that varies with time and age, contributes to the difficulties in managing MS. There is a need for development of therapies which modulate the immune system without side effects, address the inflammation and help in neuro-protection and remyelination [2, 3].

As gut dysbiosis has been pointed to as a significant factor underlying the disease pathogenesis and progression of MS with the severity of the disease having shown correlation with the abundance of microorganisms such as *Proteobacteria, Blautia, Dorea*, etc., and decrease in microorganisms such as *Parabacteroides, Bacteroides, Prevotella, Adlercreutzia* [4,5], a beta glucan from the N-163 strain of the black yeast *Aureobasidium pullulans* (Commercial name: Neu-REFIX) which has been reported to have beneficial reconstitution of the gut microbiome in several pre-clinical and clinical studies with increase in the beneficial microorganisms and decrease in harmful microorganisms [6,7] that are relevant to MS, could be worth as a potential

DMT. The gut microbiome reconstitution in these studies with this Neu-REFIX beta glucan, has been correlated with improvement in clinical parameters such as decrease in inflammation apart from positive effects on behaviour, sleep, alpha-synuclein and melatonin levels [8,9].

In another study in human healthy volunteers, this N-163 strain produced beta-glucan mitigated inflammation, as evidenced by decreases in anti-inflammatory markers like CD11b, serum ferritin, galectin-3, and fibrinogen [10]. It has also resulted in positive immuno-modulation through a reduction in the neutrophil-to-lymphocyte ratio (NLR) and an increase in the lymphocyte-to-CRP ratio (LCR) and leukocyte-to-CRP ratio (LeCR) [11]. In a study on mice it could reduce the inflammatory cascades linked to lipotoxicity [12]. Another investigation using an animal model of non-alcoholic steatohepatitis (NASH) revealed accumulation of F4/80+ cells, which are macrophages linked to inflammation, and a reduction in liver inflammation [13]. In patients with COVID-19, this N-163 strain produced beta glucan along with another AFO-202 strain produced beta glucan has been shown to decrease acute cytokine storm related factors such as CRP, IL-6, D-Dimer, Ferritin apart from decrease in NLR with increase in LCR and LeCR in a duration of 15 and 30 days [14,15]. Considering the safety of the referred B-glucans and their reported reconstitution of gut microbiome [4,5], in the present study we have evaluated the safety and efficacy of the N-163 strain of A. pullulans-produced beta 1-3,1-,6 glucan in patients with MS, in terms of clinical parameters along with evaluation of the biomarkers of relevance to inflammation and auto-immunity.

## Methods

This trial involved patients with MS and was an open label, prospective, non-randomised, non-comparative single arm clinical study. The study was of 60 days duration.

Twenty patients were screened while 18 patients were found eligible to participate as per the inclusion criteria given below.

### Inclusion criteria

1. Adults aged above 21 years of age; all genders inclusive; with diagnosis/history of MS
2. Subjects who have been on the same treatment regimen for a minimum of three months prior to enrolment and are willing to not make major changes to their standard treatment regimen until the end of the treatment period.
3. Subject/LAR who is willing to give written informed consent for participation, able to comprehend and understand the responsibilities during treatment period.
4. Subjects who are willing not to participate in any other clinical trial during participation in the current trial.

### Exclusion Criteria

1. Subjects with history that suggests possible allergic reaction to the key constituents of the investigational product.
2. Subjects who have difficulty in swallowing or any condition that makes per oral medication difficult or impossible
3. Subjects who have undergone major surgical procedure 4 weeks prior to randomisation.
4. Subjects who are on anti-depressants, anti-psychotics or presenting in psychiatric condition that would interfere with the parameters of the clinical study.
5. Subjects with CKD or other diseases that impair normal kidney function.
6. Subjects with known history of clinically significant endocrine, gastrointestinal, cardiovascular, hematological, hepatic, immunological, renal, respiratory, or genitourinary abnormalities or diseases; except those that are considered etiology or co-morbid to the study indication.
7. Females who are pregnant or lactating or planning to become pregnant during the study period.
8. Subjects who are currently participating or have participated in a clinical trial upto 90 days prior to randomisation
9. Subjects, who in the opinion of the investigator are unsuitable for enrolment.

## Treatment regimen

The treatment regimen followed by the patients prior to the study was heterogeneous both in the choice of the medicines and the treatment duration but majorly consisted of steroids, muscle relaxants and in some of the patients, interferon beta-1b (IFNB) and immunomodulators such as alemtuzumab, fingolimod, or natalizumab.

The patients consumed two sachets of N-163 strain of *A*.*pullulans* produced beta-glucan (each sachet of 8g gel, containing 48 mg of active ingredient)(commercial name: Neu-REFIX), supplied by GN Corporation Co. Ltd., Japan for 60 days in addition to their above mentioned standard treatment regimen.

### Evaluations performed

The following evaluations were performed at Day 1 and Day 60 of the trial

1. Blood Levels of Inflammatory and immunomodulatory parameters such as IL-6 CRP, ESR, CD56+ve cells, CD16+ ve cells, CD4+ve cells, CD8+ve cells, CD3+ve cells and CD19+ve cells
2. Kurtzke Expanded Disability Status Scale (EDSS) score

### Safety

Aside from vital sign monitoring, physical and neurological examinations, clinical laboratory tests and concurrent medications, safety outcome measures also included the frequency and type of adverse events (AEs), serious AEs, discontinuations for AEs, and vital sign measurements.

### Statistical analysis

This single-arm noncomparative study’s analysis was exploratory. There were no explicit hypotheses tested. The intention-to-treat (ITT) population, which consisted of all recruited patients (n=18) who received the N-163 beta glucan, including those who withdrew early and underwent no evaluations, was used for the efficacy analyses. All patients from the ITT sample that were not patients with both missing screening and baseline EDSS scores were referred to as the modified ITT population.

All data were analysed using Excel statistics package analysis (Microsoft Office Excel) and Origin2021b software. All 27 randomized participants were included in safety analysis sets. All statistical tests were performed at a significance level of 0.05 without correction for multiple comparisons or multiple outcomes. Non-parametric tests such as Mann-Whitney U test and Kruskal-Wallis tests were used.

## Results

Eight females and four males participated in the study. The mean age was 37.2 years. All the participants in this study were of Indian descent. The anthropometric measurements (height, weight, and BMI) showed no significant changes between the baseline and study’s conclusion.

The values are represented as mean ± standard deviation. There was decrease in CD16 + ve cells’ count from 1.37 ± 1.0 to 1.25 ± 1.75 (t = 1.34, p = 0.104 > 0.05). There was decrease in CD4 +ve cells 46.13 ± 14.51 to 45.82 ± 7.18 (t = -1.73, p = 0.944 > 0.05). There was increase in CD8 +ve cells from 29.12 ± 6.13 to 29.36 ± 7.6. There was a slight decrease in CD3 +ve cells from 78.62 ± 6.96 to 78.55 ± 7.73 (t = -3.42, p = 0.997) > 0.05). There was a decrease in CD19+ ve cells from 12.58 ± 6.30 to 10.54 ± 7.25 (t = -1.75,p =0.946 > 0.05). The improvement in CD56 from baseline to end of study in an increase of 1545.21% (t = 1. 73, p = 0.056 > 0.05) (Figure 1). There was a decrease in IL-6 from 27.74 ± 64.7 to 9.67 ± 5.63 (t = 1.45, p = 0.912 > 0.05) (Figure 2). There was an increase in LCR from 36.69± 32.6 to 90.64 ± 155.8 and LeCR from 7197.63 ± 5342.6 to 19266.9 ± 35619.0. There was a decrease in NLR from 3.08 ± 1.84 to 2.66 ± 1.05 (Figure 2). The EDSS score (n=11) improved by 0.5 in two patients, 1 in one patient, worsened by 0.5 in 1 patient while remaining stable in all other patients (Figure 3).

**Figure 1:**
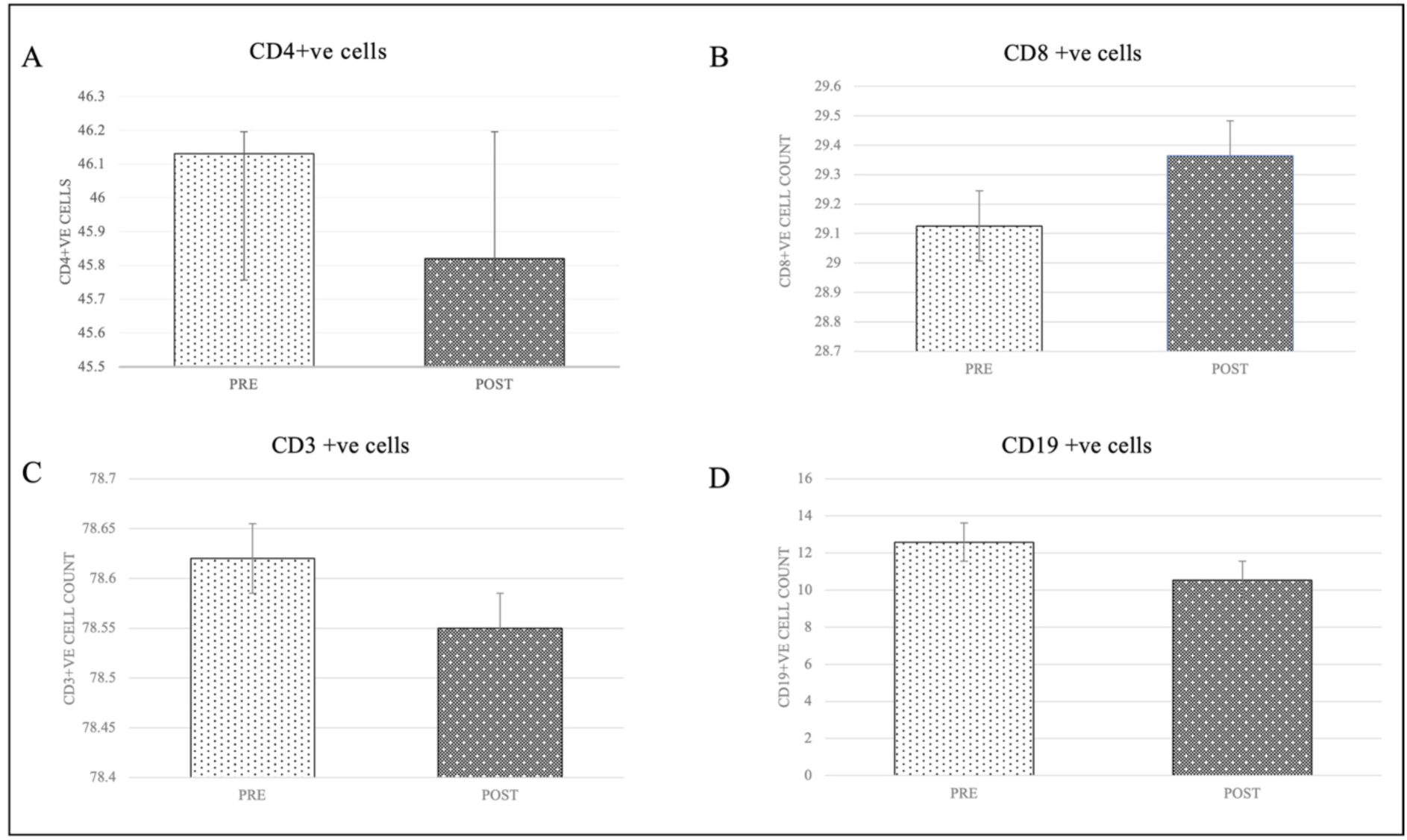
Beneficial improvement in immune-modulation associated parameters A. Cd4 +ve cell count; B. CD8+ve cell count; C. CD3+ve cell count and D. CD19+ve cell count in MS patients pre-and post-N-163 strain of A.*pullulans* produced B-Glucan

**Figure 2:**
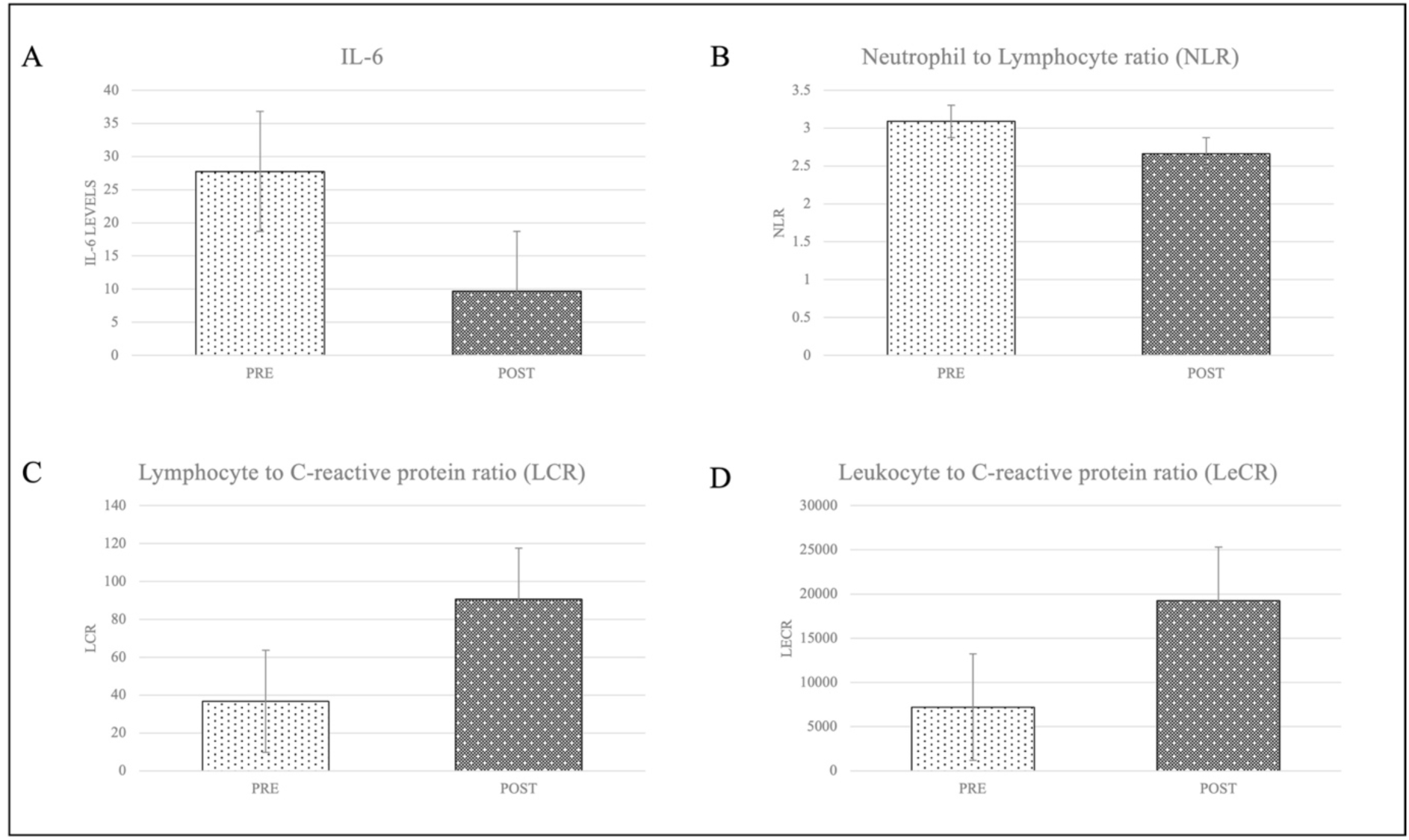
Beneficial improvement in inflammation associated parameters A. IL-6, B. neutrophil to Lymphocyte ratio (NLR), C. Lymphocyte to C-reactive protein ratio (LCR) and D. Leukocyte to CRP ratio (LeCR) in MS patients pre-and post-N-163 strain of *A.pullulans* produced B-Glucan

**Figure 3:**
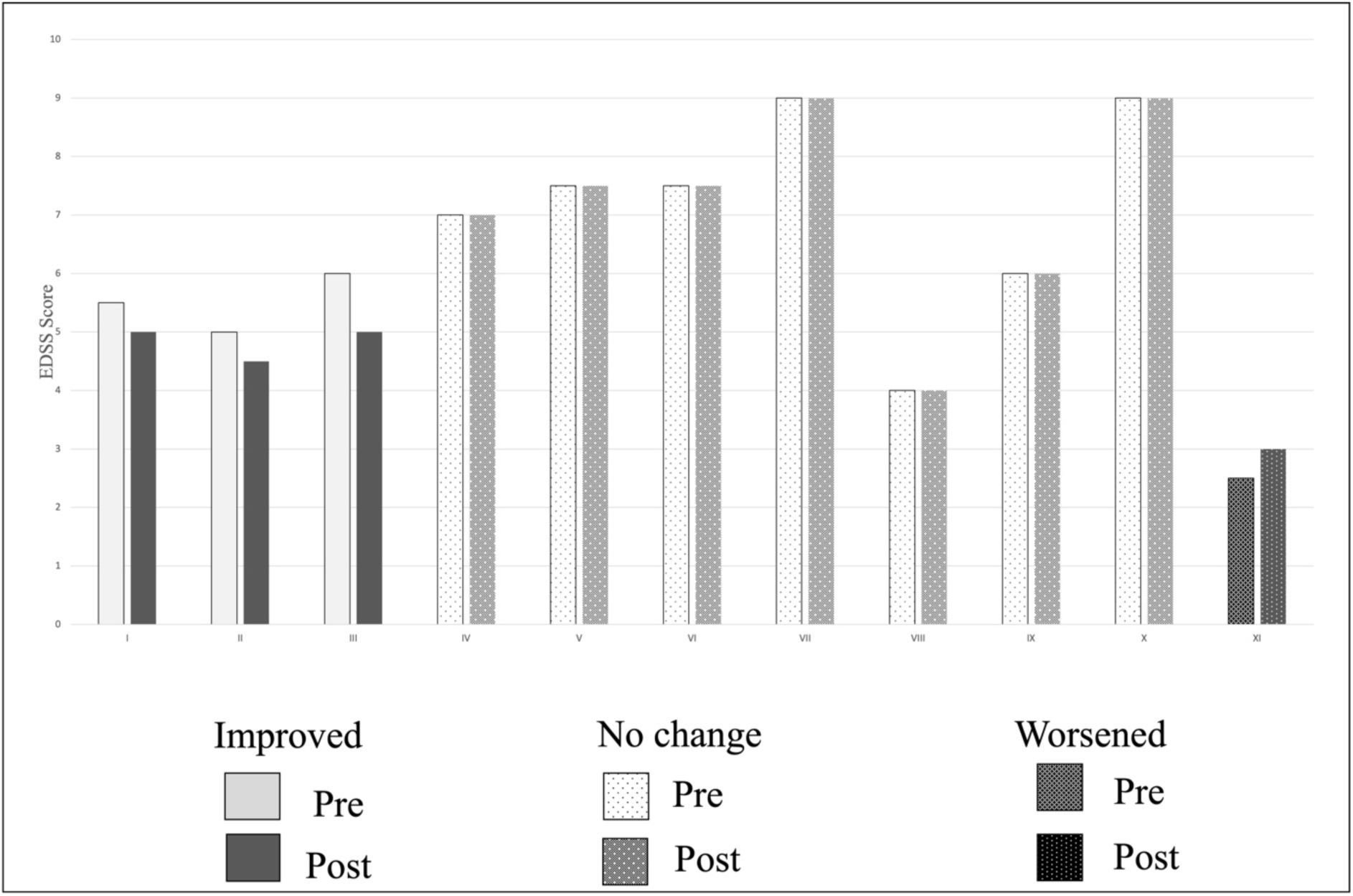
Expanded Disability Status Scale (EDSS)in MS patients pre-and post-N-163 strain of *A*.*pullulans* produced B-Glucan

The vital signs that were measured at the beginning of the trial and at its conclusion showed no significant change.

A total of five adverse events were reported during the study but none of them were related to the investigational product (Neu-REFIX B-glucan). The adverse effects reported were one case of lower back pain, one case of vomiting, one case of fever, one case of excessive tiredness and one case of skin rashes. Thus N-163 Nichi Glucan is proven to be safe to be consumption for treatment. All these AE outcomes were documented as being resolved before to the study’s conclusion.

## Discussion

A chronic inflammatory condition of the central nervous system (CNS), multiple sclerosis (MS) primarily affects young individuals [16]. The most widely accepted working theory on the pathophysiology of MS postulates that autoreactive T cells escape from clonal deletion in the thymus and a defective regulatory system in the periphery before being (re-)activated in lymphoid tissues by as of yet unidentified triggers. Finally, these autoreactive cells enter the central nervous system (CNS) through the blood-brain barrier, causing myelin and oligodendrocyte damage that leads to gliosis, neuro-axonal damage, and inflammation. The course of the disease is accelerated in the latter stages by compartmentalised inflammation that fuels ongoing CNS inflammatory and degenerative processes [16]. According to another hypothesis called “inside-out hypothesis,” CNS antigens are released as a result of the early loss of oligodendrocytes and myelin due to some etiology sets up immunological responses against myelin components, which ultimately lead to neuroinflammation [17]. The cause of MS is still unknown, and no recognised triggers have been found for either a primary inflammatory illness beginning or a primary oligo-dendro-glial pathology followed by inflammation [18].

Patients with secondary progressive MS (SP-MS) and RR-MS showed decreased CCR9 functioning [19,20] in earlier reports from the literature. CD4+ T cells increase the expression of CCR9+ on T cells. Therefore decrease of CCR9+ CD4+ T cells in peripheral blood caused by the inhibition of the CCR9-CCL25 interaction has been reported to be beneficial. Studies of MS have demonstrated the central contribution of CD4+ T lymphocytes in the pathogenesis of the disease [19]. In the present study there was decrease in CD4+ve cells.

NLR, LCR and LeCR are established inflammation markers that reflect systemic inflammatory response. Increased NLR levels and low LCR, LeCR levels are reflective of an increased inflammatory process [20]. Especially in COVID, these markers have been used to assess the level of systemic inflammation and increased NLR, decreased LCR, LeCR associated with poor disease prognosis [20]. Several different inflammatory blood biomarkers, have been investigated in the search for an accessible biomarker useful for diagnosing MS disease and predicting disease course of which these markers NLR, LCR and LeCR are considered potential ones. NLR has been connected to autoimmune conditions like chronic inflammatory bowel illness and rheumatoid arthritis and also earlier studies have pointed to a link between elevated NLR and MS [20]. In the current study, after consumption of N-163 strain of *A*.*pullulans* produced B-Glucan, there was a decrease in NLR and increase in LCR and LeCR indicative of beneficial management or reduction of systemic inflammation which correlated with the beneficial clinical outcome in other parameters. Thus, investigating the role of such simple biomarkers in MS patients may help in understanding the pathogenesis, progression, remission and therapeutic efficacy of management strategies for MS.

Before the development of interferon beta in the early 1990s, there were no known treatments for multiple sclerosis that could alter the course of the disease. Before oral medicines were ultimately established, MS treatments primarily consisted of injection and infusion drugs for almost two decades. The first-line MS injectable medications are interferons. The possibility of producing neutralising antibodies raises the risk of injection-site responses, flu-like symptoms, and liver dysfunction, which reduces their efficacy. As a result, fresh medications dimethyl fumarate (DMF) were authorised for the treatment of MS [21] but they have side effects including cardiovascular and hepatic effects [22].

The range of MS treatments keeps expanding, yet treating MS is still quite difficult, especially in a safe manner.

Also, there is still a need for extremely efficient treatments that can be applied at any stage of the illness process preferably without or with minimal side effects. One of the important clues in clinical evaluations has pointed to the implications of the gut microbiome, correlating with the severity of the disease [23]. An increase in specific bacterial population has shown to contribute to pro-inflammatory responses in MS patients [4,5]. Earlier studies of the N-163 beta-glucan modulating the gut microbiome towards eubiosis apart from contributing to anti-inflammatory activities via the gut-brain-immune axis, indicates that this N-163 beta-glucan via multiple mechanisms including modulation of gut microbiota has contributed to a decrease in inflammation associated parameters in the present study.

Confirmed Disability Improvement (CDI) with a specific decline in the EDSS score, confirmed over a specific time period has been reported to be a key indicator of the restoration of function and therapies that lead to CDI have shown to result in better prognosis and quality of life in MS [24]. In the current study, correlation of the above biomarkers to EDSS improvement, though modest is yet significant as the duration of the trial is very short (60 days) compared to CDI which has been reported majorly in trials of at least 6 months. This outcome makes this safe orally consumable being safe orally consumable Neu-REFIX beta-glucan a potential CDI producing disease modifying drug adjuvant.

However, the limited sample size and the briefer treatment period are two major limitations of this clinical investigation. Further, larger multicentric study on the influence of the gut microbiome and the clinical parameters of relevance in the disease pathogenesis can throw light on disease modifying therapeutics that will have potential use in MS. While undertaking such studies imaging of the brain should also be included which has not been undertaken in this pilot study

## Conclusion

This pilot clinical study demonstrates the safety of the N-163 beta glucan food supplement and its efficacy in improving EDSS in three of the MS patients is a significant progress considering the current scenario where there is no definitive remedy for MS. Correlation of the positive effects on control of systemic inflammation reflected by biomarkers such as IL-6, CD4+ cells, LCR, LecR and NLR in a short duration of 60 days with a beneficial clinical outcome further strengthens the efficacy component and make us recommend larger multicentric studies of longer duration to validate the potentials of Neu-REFIX as a safe drug adjuvant in MS. Identifying the processes responsible for these positive outcomes including research on the gut microbiota composition pre-and post N-163 beta glucan intervention and also evaluation of simple immunological parameters like NLR, LCR, LeCR that might help gain an insight into the dynamically changing complex immune system in diseases such as MS should be undertaken in future studies.

## Data Availability

All data produced in the present work are contained in the manuscript

## Acknowledgements

The authors thank

1. The Government of Japan and the Prefectural Government of Yamanashi for a special loan and M/s Yamanashi Chuo Bank for processing the transactions.
2. Ms. Ann Gonsalvez and members of Multiple Sclerosis Society of India (MSSI)-Chennai Chapter for their coordination with the participants of the study.
3. Dr. Malcolm Jeyaraj, Neurologist for his clinical inputs and evaluation of the patients.
4. Dr. Ragaroobine, Mr. Rajmohan from Nichi-In Centre for Regenerative Medicine (NCRM) for their assistance with data collection.
5. Mr. Masato Onaka, Mr. Yasushi Onaka of Sophy Inc, Kochi, Japan for technical advice.
6. Ms. Yoshiko Amikura and staff of GN Corporation Co Ltd, Japan for their liaison assistance with the conduct of the study.
7. Loyola-ICAM College of Engineering and Technology (LICET) for their support to our research work.

